# Deep learning-based precision phenotyping of spine curvature identifies novel genetic risk loci for scoliosis in the UK Biobank

**DOI:** 10.1101/2025.09.03.25335040

**Authors:** Michael Zeosky, Eucharist Kun, Siddharth Reddy, Devansh Pandey, Liaoyi Xu, Joyce Y. Wang, Chenfei Li, Ryan S. Gray, Carol A. Wise, Nao Otomo, Vagheesh M. Narasimhan

**Author notes:** Contributed equally to this work (Michael Zeosky, Eucharist Kun). To whom correspondence should be addressed **Corresponding authors** Correspondence to Vagheesh M. Narasimhan.

## Abstract

Scoliosis is the most common developmental spinal deformity, but its genetic underpinnings remain only partially understood. To enhance the identification of scoliosis-related loci, we utilized whole body dual energy X-ray absorptiometry (DXA) scans from 57,887 individuals in the UK Biobank (UKB), and quantified spine curvature by applying deep learning models to segment then landmark vertebrae to measure the cumulative horizontal displacement of the spine from a central axis. On a subset of 120 individuals, our automated image-derived curvature measurements showed a correlation 0.92 with clinical Cobb angle assessments, supporting their validity as a proxy for scoliosis severity. To connect spinal curvature with its genetic basis we conducted a genome-wide association study (GWAS). Our quantitative imaging phenotype allowed us to identify 2 novel loci associated with scoliosis in a European population not seen in previous GWAS. These loci are in the gene *SEM1/SHFM1* as well as on a lncRNA on chr 3 that is downstream of *EDEM1* and upstream of *GRM7.* Genetic correlation analysis revealed significant overlap between our image-based GWAS and ICD-10 based GWAS in both the UKB and Biobank of Japan. We also showed that our quantitative GWAS had more statistical power to identify new loci than a case-control dataset with an order of magnitude larger sample size. Increased spine curvature was also associated with increased leg length discrepancy, reduced muscle strength and decreased bone density, and increased incidence of knee but not hip osteoarthritis. Our results illustrate the potential of using quantitative imaging phenotypes to uncover genetic associations that are challenging to capture with medical records alone and identify new loci for functional follow-up.

## Introduction

Scoliosis is a complex musculoskeletal disorder characterized by an abnormal lateral curvature of the spine^1,2^. This disorder manifests from a variety of causes with a small proportion caused by rare Mendelian mutations or congenital vertebral anomalies that present from birth. However, the most common forms of scoliosis are idiopathic forms, which can manifest in early childhood, adolescence, or adulthood^3,4^. Untreated scoliosis can lead to a range of complications, including chronic pain and increased risk of fractures, significantly impacting the quality of life for affected individuals. Given that scoliosis is often identified only after progression to noticeable spinal deformity, understanding the genetic basis of scoliosis is critical for advancing treatment strategies and enabling earlier identification of the condition. Despite the multifactorial nature of scoliosis, encompassing both genetic and environmental contributions^5^, its underlying genetic factors remain poorly understood. Identifying these also has increases the potential for the identification of therapeutic targets to prevent progression and improve patient outcomes.

Genome-wide association studies (GWAS) have provided valuable insights into the genetic architecture of scoliosis. Notably, several GWAS conducted in Japanese populations have identified susceptibility loci associated with adolescent idiopathic scoliosis (AIS) using binary health record data from the Biobank of Japan^6,7^. A recent meta-analysis of these studies has expanded the total number of reported loci to 21^8^. Another meta-analysis of AIS GWAS across multiple populations replicated many of these reported loci as well as discovered a locus that had sexually dimorphic effects for idiopathic scoliosis^9,10^. These genetic discoveries come from a mix of biobank and disease-specific cohorts, however, no GWAS has successfully identified genome wide significant loci from purely cohort studies such as the UK Biobank (UKB) for scoliosis, as the total number of cases of individuals coded as having scoliosis is only 4,851, or less than 1%.

A primary limitation of unselected cohort studies is the use of ICD-10 records to determine disease status. In the clinic, the severity of scoliosis is often classified using the Cobb angle, made by drawing lines along the upper and lower endplates of the most tilted vertebrae in a spinal curve and measuring the angle where these lines intersect^1^. However, this nuance is often lost in the medical record and simply classified as either having scoliosis or not. In addition, these binary case-control assessments derived from health records typically only include individuals that have extreme severe disease, thereby greatly limiting the number of cases in datasets like population biobanks. Furthermore, coding individuals as cases in the medical record could be dependent on clinician experience, the severity they consider constitutes a diagnosis, or the patient’s access to care or insurance provided^11^. Recent advancements in deep learning and computer vision provide an opportunity to overcome these limitations by directly assessing disease phenotypes from radiographic images, thus generating quantitative image-derived phenotypes (IDPs). In musculoskeletal diseases, imaging has been the primary method of diagnosis and tracking of disease progression in the clinic for decades^1^. Therefore, deep learning has the potential to directly capture nuanced disease severity in an accurate and replicable way that can improve gene discovery for such disorders. In previous work, we have shown that utilizing deep learning models for phenotyping of radiographic images increases the power of gene discovery of knee osteoarthritis^11^, and others have also applied such approaches to understand the genetic basis of hip joint space^12^. However, in scoliosis, though methods have been designed to automatically measure spinal curvature^13^, these have not been extended to study the genetic drivers of the disease. In this study, we utilize deep learning and novel image analysis methods to produce quantitative image-derived phenotypes associated with spine curvature - a natural continuous measure of scoliosis severity. By training an image segmentation model to isolate the spine from existing dual energy X-ray absorptiometry (DXA) images and further developing a method to quantify the cumulative lateral curvature of the spine on ~60,000 individuals, we provide a quantitative measure of the disease over and beyond the ICD-10 scoliosis diagnosis health record data (M41)^11^. While ascertainment of patients with scoliosis is typically carried out in adolescents, like those utilized in the previous GWAS studies, this analysis examines spine curvature using DXA scans of adult individuals encompassing the different periods in which scoliosis can manifest.

## Results

### Dataset and quality control of DXA imaging and genetic data

To study the genetic basis of spine curvature, we jointly analyzed paired DXA imaging and imputed genome sequence data of 69,981 individuals in the UKB^14^. We first restricted the dataset to individuals of white British ancestry. Next, as the bulk imaging data from the UKB consisted of DXA images that reflect scans of different body parts, we used a deep-learning approach to subset the imaging dataset to only full body transparent DXA scans^15,16^. We then removed individuals that had outlier image resolutions or poor quality DXA scans, and padded images to a standard size for processing. For the genetic data, we applied standard variant and sample quality control (QC) and analyzed 7.2 million common biallelic SNPs with minor allele frequency greater than 0.1%. Post quality control, we were left with combined imaging and genetic data for a total of 57,887 individuals aged between 46 and 81 with a mean age of 64. The ratio of males to females was 50.2% to 49.8% in this analysis, consistent with the overall distribution in the UKB.

### A deep learning model for image segmentation and quantification of spine curvature

To perform automated measurement of spinal curvature we used a training dataset of 63 images (50 training, 13 validation, **Supplementary Table 1**) (**Fig. 1a**). On each of these images, we manually outlined the spine from the top vertebrae to the bottom. We then trained a deep-learning model based on the U-Net architecture^17,18^ with a 34-layer ResNet encoder^19^ to perform semantic segmentation of the spine at pixel-level resolution (**Fig. 1b**). We then applied this model to all the 57,887 individual full body transparent images and obtained spine segmentation masks for each DXA image. To measure spine curvature, we obtained landmarks at 20 different positions along the spine mask reflecting the pixel centroids at each segment of the spine (**Fig. 1c**). Then taking a line between the top landmark and the bottom landmark of the mask, we rotated the spine image such that the line was in a vertical orientation (**Fig. 1d**). This then allowed us to calculate the overall cumulative spine curvature simply by examining the horizontal displacement between subsequent landmarks. Our spine curvature phenotype was therefore quantified as the sum of the absolute differences between x-coordinates of adjacent centroids along the spine, providing a continuous measure of lateral deviation (**Fig. 1e**). We normalized these measurements across images by converting pixel lengths to mm, which we accomplished by multiplying the pixel lengths by the pixel to mm scaling reported for each image provided by the scanner.

**Figure 1.**
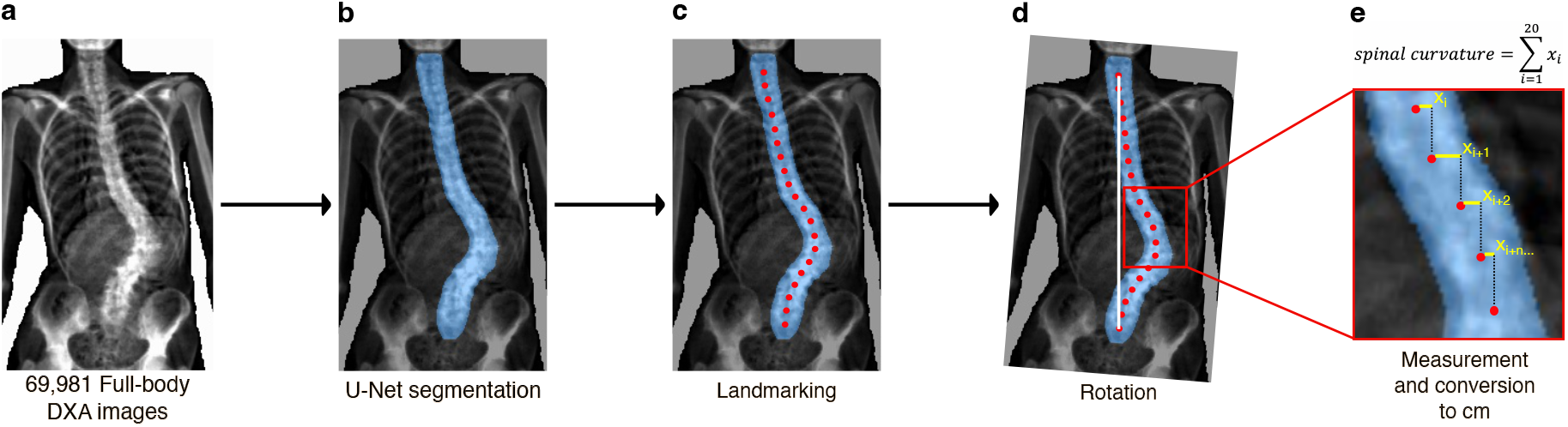
Deep learning segmentation and quantification of spinal curvature. **a.**Original DXA scan converted from DICOM format to high resolution JPEG image for downstream analysis. **b**. Deep-learning-based segmentation of spine from original image using a U-net architecture^17,18^ with a 34-layer ResNet encoder^19^. **c**. Automatic identification of key landmarks throughout the spine through the calculation of evenly spaced centroids. **d**. Rotation of spine to a vertical orientation **e**. Measurement of overall spine curvature as the sum of absolute horizontal displacement between landmarks (marked in yellow).

### Evaluation of the spine curvature measurements

We evaluated the performance of the segmentation model in several ways. First, the set accuracy - the pixel correspondence between the labeled masks and annotation of the trained model on validation data - was 0.99. Second, the correlation between images taken of the same person across two imaging visits was 0.68, despite changes in image resolution, scanner, technician, and back posture of each subject (**Fig. 2a**). We do not expect to see 100% concordance across these replicates as differences in spinal curvature can arise due to actual differences in posture or due to scoliosis. Third, we examined the correlation between Cobb angle as assessed manually by certificated orthopedic surgeons and our spinal curvature measurement in 124 individuals, which exhibited a significant Pearson correlation of 0.92. Additionally, from these 124 individuals, we examined differences in spinal curvature based on scoliosis diagnosis from Cobb angle measurement. Individuals with a Cobb angle exceeding 10 degrees, which is the typical cutoff for scoliosis diagnosis, had a statistically significant higher spinal curvature than individuals with a Cobb angle of less than 10 degrees (t-test, *P* < 3.4 × 10^−8^). We also confirmed this by examining the relationship between the spinal curvature phenotype and scoliosis status as captured in the ICD-10 code M41 case–control data^14^ (**Fig. 2b**). As expected, the spinal curvature phenotype was significantly higher in cases compared to controls (t-test, *P* < 2.2 × 10^−16^). Finally, we examined the relationships between the spine curvature phenotype and age—which is also known to be highly associated with spinal compression and curvature. Again, as expected, we found that the average spinal curvature phenotype increased significantly with age (linear regression, ß = 0.019, *P* < 0.0001) (**Fig. 2c**). Taken together, these suggest that our spine curvature phenotype was replicable, was highly correlated with Cobb angle – the clinical measure of scoliosis – and captured a certain proportion of actual inherent deviation in the vertebral column and not just deviation due to posture at the time the image was taken.

**Figure 2.**
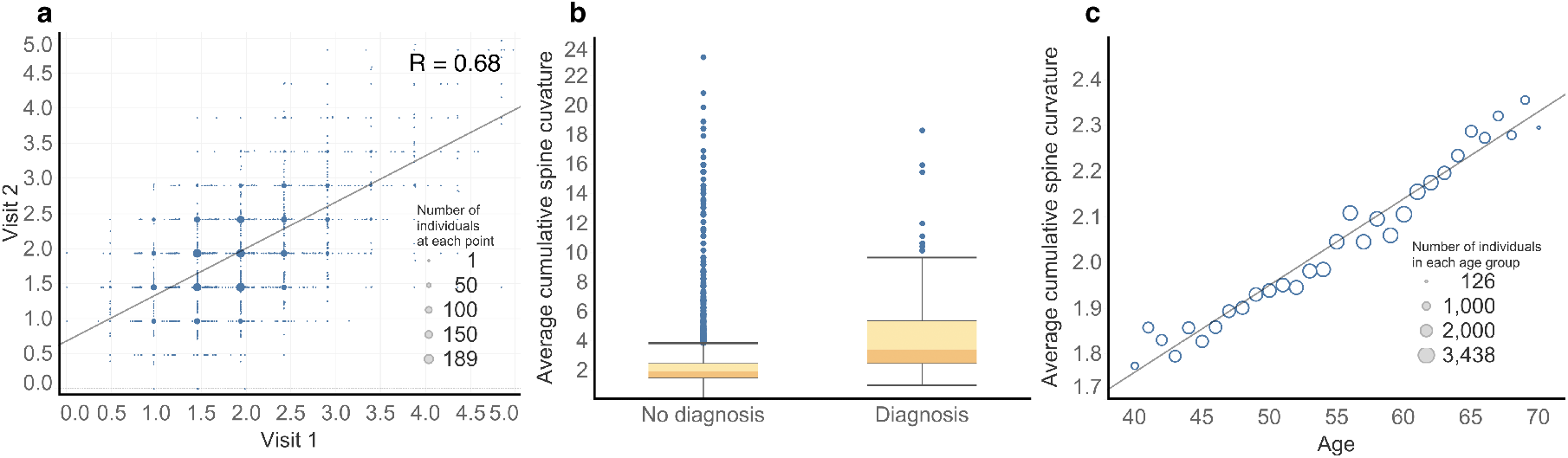
Evaluation of cumulative spine curvature across visits, ICD-10 scoliosis diagnosis status, and age. **a** Correlation of spine curvature between two visits for individuals who were imaged multiple times, (R=0.68, *P* <0.0001). **b** Comparison of cumulative spine curvature between individuals with and without a recorded ICD-10 M41 Scoliosis diagnosis. **c**. Analysis of the relationship between average cumulative spine curvature and age, with circle size correlating to the number of individuals in that age group, (beta = 0.019, *P* < 0.0001).

### Genetic associations of image derived spine curvature

Having obtained normalized spine curvature measurements on each individual, we linked this phenotype to its genetic basis. We carried out a common variant GWAS of our spine curvature phenotype using PLINK^20^ on 57,887 white British individuals adjusting for age, sex, and the first 20 genetic principal components. Using the summary statistics of this GWAS, the estimated heritability for spine curvature using Linkage Disequilibrium Score Regression (LDSC)^21^ was 0.071 ± 0.01. In total, our GWAS identified 3 genome-wide significant loci associated with scoliosis at a threshold of *P* = 5 × 10^-8^ while a GWAS performed using the UKB ICD-10 scoliosis diagnosis data on the non-imaged individuals found no genome-wide significant loci (**Fig. 3a**-**3b**). One of the SNPs is in a region upstream of the gene *PAX1* that has previously been implicated in other scoliosis GWAS and functionally validated in Zebrafish models^8,9^. Deletion of this locus has also been shown to result in tail abnormalities in female mice^22^. However, we identified two novel loci, never previously reported by other studies of scoliosis or related traits in the GWAS catalog. The first is near the gene *SEM1/SHFM1*. Deletions in this region have been shown to be associated with the rare limb developmental disorder, split hand/split foot malformation type 1^23^. The second locus is on chr 3 and is in a lncRNA that is downstream of *EDEM1* and upstream of *GRM7.*

**Figure 3.**
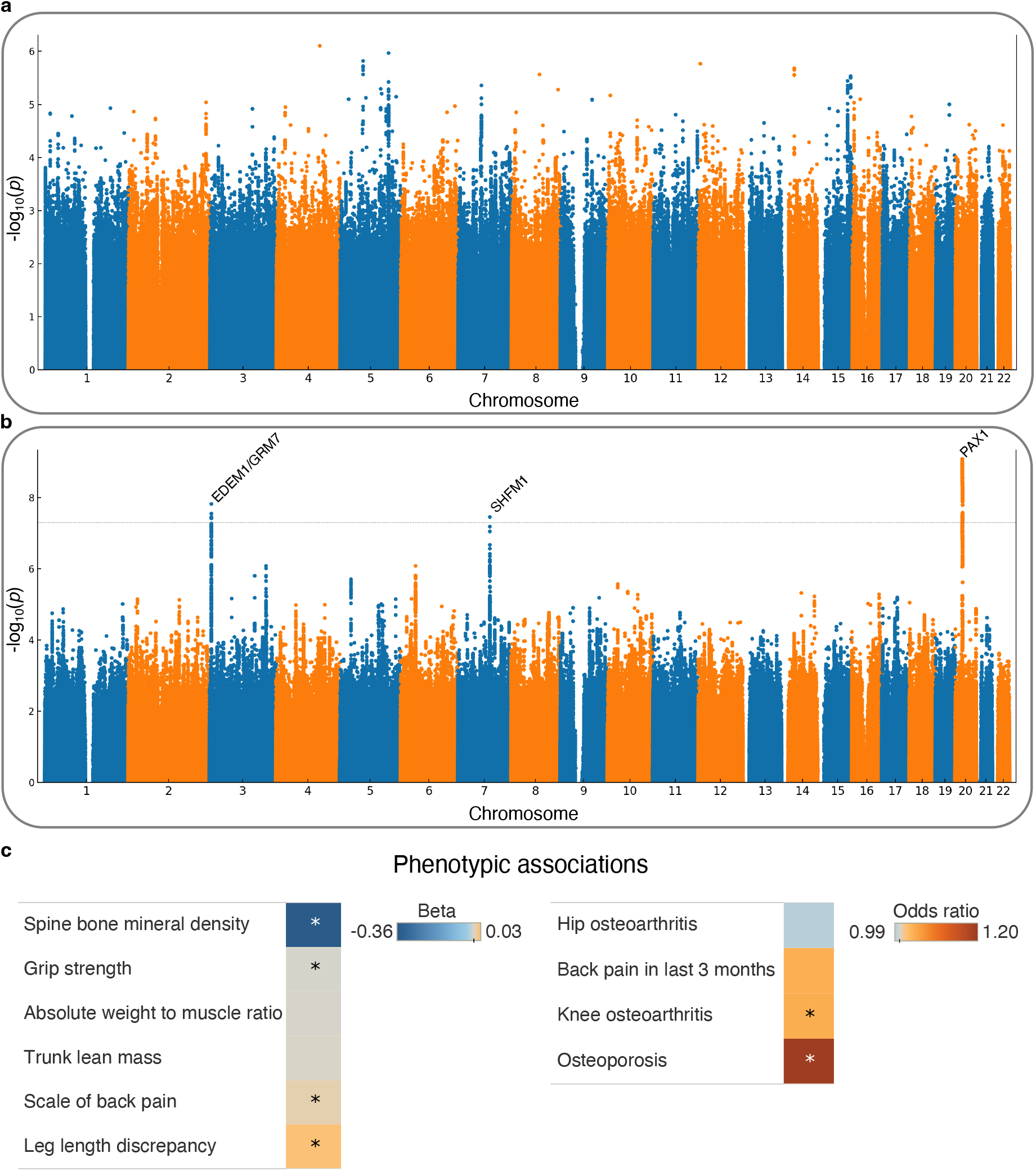
GWAS and phenotype associations **a.**Manhattan plot for GWAS performed using ICD-10 scoliosis code alone. **b**. Manhattan plot for GWAS performed using deep learning derived spinal curvature measurement. **c.** Regressions of bone and muscle phenotypes with spinal curvature. Linear and logistic regressions are shown separately. Asterisks denote significant phenotypes that have FDR-adjusted p-values < 0.05.

### Deep learning-based phenotyping improves power for discovering scoliosis associated loci

Next, we assessed the correspondence between our GWAS to other GWASs that had been performed on traditionally measured scoliosis phenotypes obtained from the electronic healthcare record. To do so, we conducted genetic correlation analyses with two case-control GWAS datasets. First, we utilized the UKB ICD-10 scoliosis diagnosis GWAS that we had previously carried out on the non-imaged individuals. The genetic correlation between this binary GWAS and our image based GWAS was 0.46 (*P* = 0.006). While not close to 1 this is within expectation for genetic correlation between a case-control GWAS and a quantitative GWAS on the same trait. In contrast, the genetic correlation between arm length, a skeletal trait that is not associated with the spine, and the UKB ICD-10 scoliosis diagnosis was not significant (*P* = 0.11). This serves as a negative control and suggests that our imaging-based phenotyping was identifying genetic loci that were highly correlated to the medical record-based diagnosis for scoliosis. Second, we examined genetic correlation with a case-control GWAS for scoliosis carried out in a Japanese population^8^ using POPCORN^24^ to adjust for differences in LD Structure between Europeans and Japanese. This demonstrated a slightly higher genetic correlation of 0.56 (*P* = 1.18 × 10^-6^). These findings suggest that our continuous, image-derived phenotypes effectively capture scoliosis-related genetic signals identified using diagnostic records while offering improved statistical power.

### Associations of scoliosis with bone and muscle density and musculoskeletal disease

We then examined the association between spine curvature and bone and muscle strength. After controlling for age and sex, we found that in line with epidemiological data, hand grip strength (a proxy for muscular strength) and spine bone density were both negatively associated with spine curvature (multivariate regression covariate p-value for grip strength: *P* = 1.4 × 10^−8^, p-value for bone mineral density: *P* < 2.2× 10^−16^) (**Fig. 3c**). We next examined the association between leg length discrepancy and scoliosis. An actual difference in bone length between the legs is thought to contribute to scoliosis, as the spine bends to accommodate the discrepancy. To examine this hypothesis, we utilized deep learning approaches to measure the difference in the length of the left and right leg. Though the difference between the length of the legs is small, only a few millimeters, we were able to replicably measure this difference in the same individual across two imaging visits taken more than two years apart. We found that spine curvature was positively associated with the absolute value of leg length discrepancy after controlling for age, sex and height (multivariate regression covariate p-value for leg length discrepancy: *P* = 2.58 × 10^−4^). We also found that individuals with increased spine curvature also self-reported back pain at higher rates, and had higher incidence of knee osteoarthritis, but not hip osteoarthritis in the ICD-10 code (multivariate regression covariate p-value for back pain: *P* = 4.57 × 10^−2^, knee OA: *P* = 4.84 × 10^−2^, hip OA: *P* = 0.82).

## Discussion

Our study demonstrates the potential of using deep learning-derived phenotypes, such as spine curvature from DXA images, to improve genetic analyses of scoliosis. By moving beyond traditional case–control phenotypes sourced from EHR records, we provide a quantitative measure of spinal curvature that captures a continuum of variation, uncovering subtle phenotypic differences associated with scoliosis risk. This quantitative approach circumvents challenges of diagnostic inconsistency and variability in clinical definitions, offering a more standardized and scalable method for phenotyping in biobank-scale settings^11^.

One major limitation of our study is our inability to differentiate adult scoliosis from adolescent idiopathic scoliosis. While adult scoliosis can occur as a continuation or exacerbation of adolescent scoliosis, other factors such as spine degeneration can also play a role in adult scoliosis. For reference, though cobb angle and spinal curvature measured in our cohort displayed a Pearson correlation of 0.92, the heritability of cobb angle measured in 4465 adolescent patients was 0.21 ± 0.10, while the heritability of our spinal curvature measurement in 57,887 adults was 0.07 ± 0.008^25^. To understand if the genetic signals we obtained were likely to be involved in adolescent scoliosis, we computed genetic correlation with the Japanese AIS cohort^8^ and observed a correlation of R_g_ = 0.56. This is in line with a previously reported genetic correlation of 0.60 between cobb angle and adolescent idiopathic scoliosis^25^. Additionally, as adolescent idiopathic scoliosis is more common in the thoracic region and adult scoliosis more commonly affects the lumbar region, we wanted to see if we could separate the genetic signals by dividing our spinal curvature assessment into two parts: the top half reflecting the first 10 points and focused more on thoracic vertebrae in our spine segmentation, and the bottom half focused on lumbar vertebrae. We then recomputed our GWAS on these two halves of the spine. We found that the genetic correlation between the adolescent cohort in the Japanese biobank and the bottom half curvature was R_g_ = 0.66 ± 0.15, while the top half was 0.48 ± 0.10. Thus, we were unable to detect significant differences using this approach. However, significantly non-zero genetic correlation suggests that our approach is capturing a significant proportion of the genetic basis of adolescent scoliosis as well. Similar to findings in other musculoskeletal conditions like knee osteoarthritis^11^, our results suggest that quantitative phenotyping offers increased statistical power for genetic studies compared to case–control approaches.

Despite its advantages such as improved consistency and scalability, our approach has limitations. The DXA-based quantitative phenotype remains a derived measure, and its precision relies on accurate segmentation and alignment. As the derived measure is not a cobb angle commonly used to analyze scoliosis severity, singular results can be difficult to interpret out of context. Similarly, while our model reduces bias inherent to health record data, any bias in the underlying imaging data or the segmentation model may propagate through the analysis pipeline. Additionally, as our GWAS focused primarily on individuals of European ancestry, the transferability of our findings to other populations requires further validation. Future efforts should prioritize extending this methodology to more diverse cohorts and exploring its applicability to other imaging modalities, such as MRI or CT scans.

We observed several associations between spine curvature and phenotypes such as leg length discrepancy and bone mineral density. While these associations reflect plausible biologically meaningful correlations such as leg length discrepancy driving the difference in spine curvature, we note that these analyses do not represent causal inference, and additional work is required to examine these effects.

Our study contributes to a growing body of evidence supporting the use of image-derived phenotypes for large-scale genetic research. By providing a proof-of-concept for the utility of quantitative phenotyping in scoliosis, our results underscore the broader potential for applying similar methodologies to other conditions where imaging plays a key diagnostic role. For example, directly measuring spinal curvature could serve as a model for rethinking disease phenotyping in biobanks and accelerating discoveries in complex traits and diseases.

## Methods

### UKB dataset and participants

All analyses were conducted using data from the UK Biobank (UKB), a prospective, population-based cohort that recruited 500,000 individuals aged 40–80 in the UK between 2006 and 2010^14^. The dataset offers extensive health records and whole-genome sequencing, allowing us to analyze 12.1 million common biallelic SNPs with a minor allele frequency greater than 0.1%. We focused on 402,000 participants who self-identified as white British and had available genetic data and continued consent as of February 22, 2022. Access to the dataset was granted under application number 65439.

### DXA imaging data

The UKB provides dual-energy X-ray absorptiometry (DXA) images collected using a Lunar iDXA instrument (GE Healthcare) in DICOM format as part of a bulk data field ID. For this study, we utilized full-body transparent DXA scans, with the original bulk download yielding 75,268 DICOM images from 70,405 unique participants. Images were assessed for poor quality or incorrect labeling through a previously developed deep learning pipeline created to classify types of DXA images and subsequently removed from analysis^15,16^. Each scan was associated with metadata detailing pixel and millimeter-based dimensions. All participants were instructed to lie flat on the scanner during imaging, resulting in non-weight-bearing images.

The DXA scans exhibited two distinct categories:

1. **Black background images**: 660–816 × 272 pixels
2. **White background images**: 930–950 × 300–382 pixels

To standardize image dimensions and control for scaling effects during deep learning, we applied padding to bring the images to consistent sizes. Black background images were padded to 816 × 288 pixels, while white background images were resized to 960 × 384 pixels. This process involved converting each DICOM image into a NumPy array and appending rows or columns of pixels as needed using NumPy^26^, SciPy^27^, and sci-kit image^28^. This resulted in 56,779 images of 816×288 and 18,489 images of 960×384. These images were then converted to high-resolution JPEGs using the pydicom library^29^ for downstream analyses.

### Image segmentation

We manually annotated 63 DXA scans for spine segmentation using the LabelMe software^30^, splitting the sample evenly between participants with and without a prior scoliosis diagnosis (ICD-10 code M41). Relevant demographic characteristics of the annotated dataset are summarized in **Table 1**. An example of the result of our manual annotation is shown in **Supplementary Fig. 1**.

**Supplementary Fig. 1.**
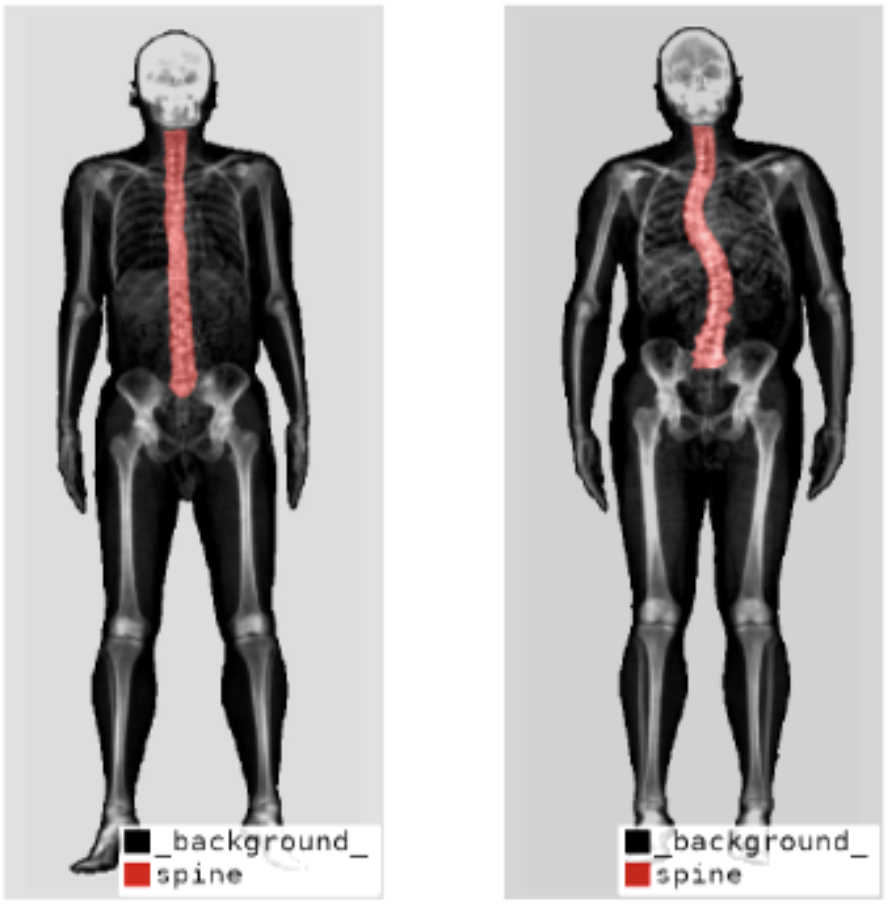
Manual labeling of the spine in two individuals shown in red.

**Supplementary Table 1.** Relevant characteristics and distribution of training data.

| Group | Characteristic | Mean (s.d.) |
| --- | --- | --- |
| **Model Training Data** | Participants, total | 63 |
|  | Age | 58 (7.24) |
| **Male** | Total (%) | 32 (50.8) |
|  | Age | 60 (7.17) |
| **Female** | Total (%) | 31 (49.2) |
|  | Age | 57 (7.04) |
| **Scoliosis** | Total (%) | 38 (60.3) |
|  | Male, total (%) | 19 (50) |
|  | Female, total (%) | 19 (50) |
|  | Age | 58 (7.06) |
| **Non-Scoliosis** | Total (%) | 25 (39.7) |
|  | Male, total (%) | 13 (52) |
|  | Female, total (%) | 12 (48) |
|  | Age | 58 (7.64) |

For segmentation we implemented a U-net architecture^17,18^ with a 34-layer ResNet^19^ encoder to perform semantic segmentation of the spine. A separate model was trained for each image size category. Both models were trained for 12 epochs with data augmentation such as rotation and zoom applied to the training data to improve generalization. To improve the robustness of the segmentation models, padded images from the other size category were subject to further image resizing during training to increase annotated sample size. Upon visual analysis of early results, we identified 224 model-predicted images for refinement and retrained the model to enhance precision. Based on human annotation, the per pixel accuracy of the segmentation was 99%. The final models were deployed on the 75,268 full-body transparent DXA images. Resulting segmentation mask files (**Supplementary Fig. 2**) were exported as JPEG images using the pydicom library^29^.

**Supplementary Fig. 2.**
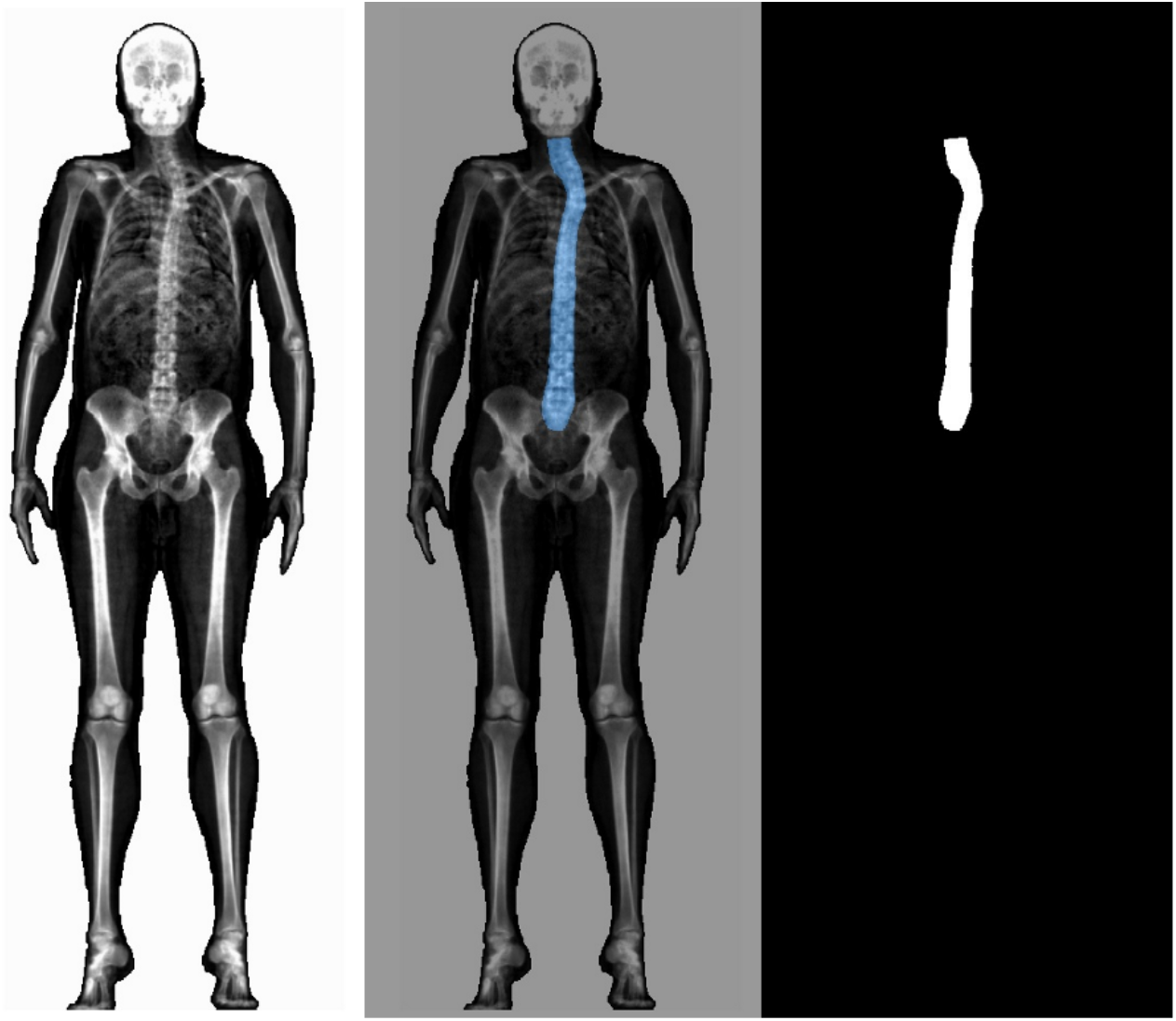
Example of the pixel mask output of our segmentation model obtained for further image processing and measurement of spine curvature.

### Spine rotation and curvature measurement

To quantify spinal curvature, we developed a custom pipeline in Python, adapted from the PlantCV package^31^, with additional modules from NumPy and OpenCV^32^. Each segmented spine mask was processed independently and converted to a binary matrix, where pixels representing the spine were set to 1, and background pixels were set to 0. Mask contours were analyzed, and a bounding box was created around the spine, whose length represented the height of the spine. This box was then divided into 20 equal-height segments to ensure consistent spatial sampling through the spine and across images. Finally, within each segment, pixels were analyzed row by row to calculate the centroid, representing the geometric center of the segment’s pixels. To assess spinal alignment, we compared the x-coordinates of the topmost and bottommost centroids. If they did not align vertically, the angle to align the points was computed and we performed a rotation using bicubic interpolation in OpenCV. The image was iteratively rotated until the centroids aligned along a vertical axis to then calculate horizontal displacement. Once the spine was aligned and centroid positions were stored, the curvature was quantified as the cumulative horizontal displacement between consecutive centroids along the spine. The sum of the absolute differences in x-coordinates between adjacent centroids provided a metric for curvature, reflecting total deviation from a straight axis (**Supplementary Fig. 3**).

**Supplementary Fig. 3.**
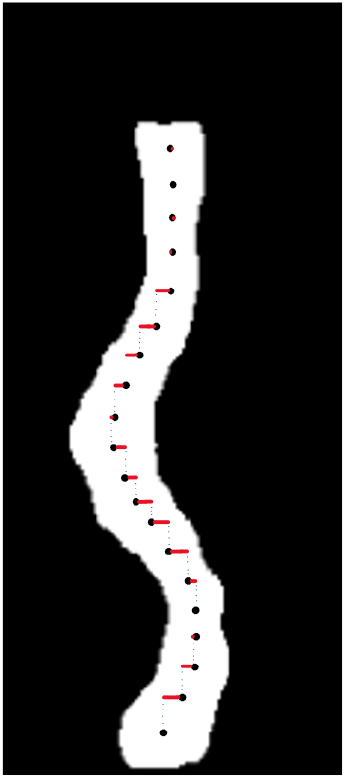
Cumulative horizontal displacement measurement To address variability in image resolution, we scaled each distance in pixels using the corresponding pixel to millimeter measurements from the metadata. These conversion ratios were validated against previous studies using an alternate measure of computing these by regressing the overall height in pixels^16^.

### Imaging data processing and quality control

To examine issues associated with our segmentation and image processing pipeline, we examined key metrics, such as top and bottom centroid positions before and after alignment, cumulative difference squared to emphasize quick changes in curvature, and pixel value counts to identify segmentation issues were exported for quality control and downstream analyses. Outliers identified based on unusual curvature values >2 standard deviations from the mean were removed. After this quality control, the final dataset consisted of 74,667 images from 69,981 individuals, ready for subsequent analyses.

To ensure the data conformed to a normal distribution, the RankNorm function in R was applied. This approach uses rank-based inverse normal transformation to standardize the data, enhancing its suitability for downstream genetic correlation analyses while minimizing the influence of outliers. This normalization is particularly useful when dealing with datasets where a few strong cases may dominate and cause other curvature data to be obscured by noise, ensuring that meaningful patterns are not lost.

### Correlation between Cobb angle measurement and image derived spinal curvature

For a set of 124 images, with clinician guidance, we measured the Cobb angle, which is the angle formed by lines drawn along the top and bottom vertebrae of the curve and typically used for scoliosis diagnosis. We used Pearson correlation to examine the association between these physician derived Cobb angle measurements and our computationally derived spinal curvature measurements. Additionally, as scoliosis patients are only diagnosed at Cobb angle measurements greater than 10 degrees, we split our cohort of 124 individuals into scoliosis and non-scoliosis groups and compared the mean spinal curvature between the two groups through a t-test.

### Genetic quality control

For our genome-wide association analyses, we focused on participants of Caucasian ancestry, self-reported as white British (FID 21000) and confirmed via genetic principal component analysis (PCA) from the UKB. We excluded individuals whose self-reported sex (FID 31) did not match their genetic sex (FID 22001), those with evidence of sex chromosome aneuploidy (FID 22019), individuals with heterozygosity or genotype missingness outliers (based on UKB’s QC procedures for DNA sample processing and genotyping, FID 22027), and those with more than nine third-degree relatives or any of unknown kinship (FID 22021). This filtering left us with a sample of 402,000 individuals. From this subset, we further restricted our analysis to participants who had undergone imaging (FID 20158) and had complete DXA measurements (FID 12254), leaving 57,887 participants for analysis. We downloaded imputed genetic data for 487,253 individuals from the UKB^14^ (covering chromosomes 1 through 22, FID 22828) and filtered this dataset according to our QC criteria using PLINK v2^20^. We excluded any duplicate SNPs (--rm-dup “exclude-all”), retained only biallelic SNPs (--snps-only “just-acgt”), limited the dataset to those with no more than two alleles (--max-alleles 2), a minor allele frequency greater than 1% (--maf 0.01), individual missingness rates below 2.5% (FID 22005), and a per-SNP missingness threshold of 5% (--maxMissingPerSnp 0.05). Following these steps, 7,204,508 SNPs remained in the imputed dataset.

### Genome-wide association studies (GWAS)

We conducted GWAS using PLINK v2^20^ with the following filtering thresholds: minor allele frequency greater than 0.01, SNP missingness below 5%, and individual missingness below 2.5%. We included the first 20 genetic principal components provided by UKB (FID 22009), sex (FID 31), and age (FID 21022) as covariates. After applying both genetic and imaging QC filters, 57,887 participants remained for the final GWAS analysis, with 7,204,508 SNPs included in each association study. SNPs identified in the GWAS were further clumped using the PLINK -- clump function with a significance threshold of 5.0 × 10^-8, a secondary significance threshold of 1.0 × 10^-4 for SNPs in linkage disequilibrium (LD), an r^2^ threshold of 0.1, a 1 Mb window, and a 20kb border around each gene. SNPs were assigned to genes using --clump-verbose and -- clump-range with the glist-hg19 reference. We annotated protein-coding genes within 250 kb of each clumped region and on the GRCH37 genome browser.

### Validation of Phenotype Consistency through Genetic Correlation Analysis

To assess the genetic similarity of scoliosis phenotypes across two populations and validate that the identified phenotype aligns with a scoliosis outcome, we carried out genetic correlation analysis. For samples of European ancestry from the UK Biobank we used Linkage Disequilibrium Score Regression^21^. For examining genetic correlation across ancestries and adjusting for LD differences, we used the Popcorn software package^24^. We examined genetic correlation between our image derived phenotypes and a GWAS from the UKB with case-control phenotypes from scoliosis diagnosis health records, and also a GWAS from participants in a Japanese population (BBJ) with case-control derived phenotypes. This analysis yielded estimates of heritability (h^2^) and genetic correlation (R_g_) between the two scoliosis phenotypes, along with associated standard errors, Z-scores, and p-values.

## Data availability

All data used for this study were obtained from the UK Biobank under application number 65439. GWAS summary statistics are currently being uploaded to the GWAS catalog and are available at this Box link: https://utexas.box.com/s/5iqlxvhul0ri9torr89lspb66thmpcdl

## Code availability

Deep-learning and image processing tools can be found at https://github.com/reb12345/Scoliosis.

## Acknowledgements

V.M.N. was supported on a grant from the Allen Discovery Center program, a Paul G. Allen Frontiers Group advised program of the Paul G. Allen Family Foundation.

## Author information

*Department of Integrative Biology, The University of Texas at Austin, Austin, TX, USA* Michael Zeosky, Eucharist Kun, Siddharth Reddy, Liaoyi Xu, Devansh Pandey, Joyce Wang, Chenfei Li, Vagheesh M. Narasimhan

*Department of Nutritional Sciences, The University of Texas at Austin, Austin, TX, USA, Department of Pediatrics, Dell Medical School, Austin, TX, USA* Ryan Grey

*Center for Musculoskeletal Research, Texas Scottish Rite Hospital for Children, Department of Orthopedic Surgery, The University of Texas Southwestern Medical Center,* Ryan Grey

*Department of Statistics and Data Science, The University of Texas at Austin, Austin, TX, USA* Vagheesh M. Narasimhan

## Contributions

M. Z. wrote the paper with input from all co-authors. E. K., S. R., L. X., D. P., J. W., C. L. and

N. O. performed the analysis. R.G., C.W. and V. M. N. supervised the analysis.

## Ethics declarations

The authors declare no competing interests

## Notes

### Competing Interest Statement

The authors have declared no competing interest.

### Funding Statement

This study was funded by a grant from the Allen Discovery Center program, a Paul G. Allen Frontiers Group advised program of the Paul G. Allen Family Foundation.

### Author Declarations

All our data comes from the UK Biobank study. The Ethics Advisory Committee of the UK Biobank gave ethical approval for our work under application ID: 65439.

### Summary of Updates

The author's name was actually spelled correctly the first time

## References

1. Janicki, J. A. & Alman, B. Scoliosis: Review of diagnosis and treatment. Paediatr Child Health 12, 771 (2007).

2. Aebi, M. The adult scoliosis. European Spine Journal 14, 925–948 (2005).

3. Janicki, J. A. & Alman, B. Scoliosis: Review of diagnosis and treatment. Paediatr Child Health 12, 771 (2007).

4. Giampietro, P. F. Genetic Aspects of Congenital and Idiopathic Scoliosis. Scientifica (Cairo) 2012, 152365 (2012).

5. Konieczny, M. R., Senyurt, H. & Krauspe, R. Epidemiology of adolescent idiopathic scoliosis. J Child Orthop 7, 3 (2012).

6. Takahashi, Y. et al. A genome-wide association study identifies common variants near LBX1 associated with adolescent idiopathic scoliosis. Nature Genetics 2011 43:12 43, 1237–1240 (2011).

7. Kou, I. et al. Genetic variants in GPR126 are associated with adolescent idiopathic scoliosis. Nature Genetics 2013 45:6 45, 676–679 (2013).

8. Kou, I. et al. Genome-wide association study identifies 14 previously unreported susceptibility loci for adolescent idiopathic scoliosis in Japanese. Nature Communications 2019 10:1 10, 1–9 (2019).

9. Sharma, S. et al. A PAX1 enhancer locus is associated with susceptibility to idiopathic scoliosis in females. Nat Commun 6, 6452 (2015).

10. Khanshour, A. M. et al. Genome-wide meta-analysis and replication studies in multiple ethnicities identify novel adolescent idiopathic scoliosis susceptibility loci. Hum Mol Genet 27, 3986–3998 (2018).

11. Flynn, B. I. et al. Deep learning based phenotyping of medical images improves power for gene discovery of complex disease. npj Digital Medicine 2023 6:1 6, 1–12 (2023).

12. Faber, B. G. et al. The identification of distinct protective and susceptibility mechanisms for hip osteoarthritis: findings from a genome-wide association study meta-analysis of minimum joint space width and Mendelian randomisation cluster analyses. EBioMedicine 95, 104759 (2023).

13. Jamaludin, A. et al. Automated measurement of size of spinal curve in population-based cohorts: Validation of a method based on total body dual energy X-ray absorptiometry scans. Bone 172, 116775 (2023).

14. Bycroft, C. et al. The UK Biobank resource with deep phenotyping and genomic data. Nature 562, 203–209 (2018).

15. Xu, L. et al. The genetic architecture of and evolutionary constraints on the human pelvic form. Science (1979) 388, (2025).

16. Kun, E. et al. The genetic architecture and evolution of the human skeletal form. Science (1979) 381, (2023).

17. Weng, W. & Zhu, X. U-Net: Convolutional Networks for Biomedical Image Segmentation. IEEE Access 9, 16591–16603 (2015).

18. Ronneberger, O., Fischer, P. & Brox, T. 2015-U-Net. ArXiv 1–8 (2015).

19. He, K., Zhang, X., Ren, S. & Sun, J. Deep Residual Learning for Image Recognition. Proceedings of the IEEE Computer Society Conference on Computer Vision and Pattern Recognition 770–778 (2015) doi:10.48550/arxiv.1512.03385.

20. Purcell, S. et al. PLINK: A Tool Set for Whole-Genome Association and Population-Based Linkage Analyses. Am J Hum Genet 81, 559 (2007).

21. Bulik-Sullivan, B. et al. LD Score regression distinguishes confounding from polygenicity in genome-wide association studies. Nat Genet 47, 291–295 (2015).

22. Ushiki, A. et al. Deletion of Pax1 scoliosis-associated regulatory elements leads to a female-biased tail abnormality. Cell Rep 43, (2024).

23. Crackower, M. A. et al. Characterization of the split hand/split foot malformation locus SHFM1 at 7q21.3-q22.1 and analysis of a candidate gene for its expression during limb development. Hum Mol Genet 5, 571–579 (1996).

24. Brown, B. C., Ye, C. J., Price, A. L. & Zaitlen, N. Transethnic Genetic-Correlation Estimates from Summary Statistics. Am J Hum Genet 99, 76 (2016).

25. Otomo, N. et al. Polygenic Risk Score of Adolescent Idiopathic Scoliosis for Potential Clinical Use. Journal of Bone and Mineral Research 36, 1481–1491 (2021).

26. Harris, C. R. et al. Array programming with NumPy. Nature 585, 357–362 (2020).

27. Virtanen, P. et al. SciPy 1.0: fundamental algorithms for scientific computing in Python. Nat Methods 17, 261–272 (2020).

28. Van Der Walt, S. et al. Scikit-image: Image processing in python. PeerJ e453 (2014) doi:10.7717/PEERJ.453/FIG-5.

29. Mason, D. et al. pydicom/pydicom: pydicom 2.3.0. (2022).

30. Wada, K. Labelme: Image Polygonal Annotation with Python.

31. Gehan, M. A. et al. PlantCV v2: Image analysis software for high-throughput plant phenotyping. PeerJ 5, e4088 (2017).

32. Bradski, G. The OpenCV Library. Preprint at (2000).

